# Bacterial superinfection pneumonia in SARS-CoV-2 respiratory failure

**DOI:** 10.1101/2021.01.12.20248588

**Authors:** Chiagozie O. Pickens, Catherine A. Gao, Michael Cuttica, Sean B. Smith, Lorenzo Pesce, Rogan Grant, Mengjia Kang, Luisa Morales-Nebreda, Avni A. Bavishi, Jason Arnold, Anna Pawlowski, Chao Qi, GR Scott Budinger, Benjamin D. Singer, Richard G. Wunderink, for the NU COVID Investigators

## Abstract

**Background:** Severe community-acquired pneumonia secondary to SARS-CoV-2 is a leading cause of death. Current guidelines recommend patients with SARS-CoV-2 pneumonia receive empirical antibiotic therapy for suspected bacterial superinfection, but little evidence supports these recommendations.

**Methods:** We obtained bronchoscopic bronchoalveolar lavage (BAL) samples from patients with SARS-CoV-2 pneumonia requiring mechanical ventilation. We analyzed BAL samples with multiplex PCR and quantitative culture to determine the prevalence of superinfecting pathogens at the time of intubation and identify episodes of ventilator-associated pneumonia (VAP) over the course of mechanical ventilation. We compared antibiotic use with guideline-recommended care.

**Results:** The 179 ventilated patients with severe SARS-CoV-2 pneumonia discharged from our hospital by June 30, 2020 were analyzed. 162 (90.5%) patients had at least one BAL procedure; 133 (74.3%) within 48 hours after intubation and 112 (62.6%) had at least one subsequent BAL during their hospitalization. A superinfecting pathogen was identified within 48 hours of intubation in 28/133 (21%) patients, most commonly methicillin-sensitive *Staphylococcus aureus* or *Streptococcus* species (21/28, 75%). BAL-based treatment reduced antibiotic use compared with guideline-recommended care. 72 patients (44.4%) developed at least one VAP episode. Only 15/72 (20.8%) of initial VAPs were attributable to multidrug-resistant pathogens. The incidence rate of VAP was 45.2/1000 ventilator days.

**Conclusions:** With use of sensitive diagnostic tools, bacterial superinfection at the time of intubation is infrequent in patients with severe SARS-CoV-2 pneumonia. Treatment based on current guidelines would result in substantial antibiotic overuse. The incidence rate of VAP in ventilated patients with SARS-CoV-2 pneumonia are higher than historically reported.

## Background

The contribution of bacterial superinfection to outcomes of severe SARS-CoV-2 pneumonia is unclear. Autopsy studies from patients with pneumonia caused by other viral pathogens, most notably influenza, suggest that bacterial pneumonia contributes to the risk of death.^1,2^ These autopsy studies included a minority of the patients with severe viral pneumonia and were skewed toward patients with severe disease and late findings. Serum biomarkers to reliably exclude bacterial superinfection in patients with severe community-acquired pneumonia (CAP) are lacking. As a result, the American Thoracic Society (ATS)/Infectious Diseases Society of America (IDSA) CAP guidelines recommend empirical antibiotic treatment for documented viral pneumonia despite weak evidence.^3^ Other influential panels have also recommended empirical antibiotics for severe COVID-19 pneumonia, appropriate for the syndrome (CAP vs. Hospital-acquired pneumonia [HAP]).^4,5^ Indeed, greater than 90% of patients with COVID-19 pneumonia receive antibiotics.^6^

Insensitivity of culture-based diagnosis of concomitant bacterial CAP is the major cause for uncertainty regarding antibiotic management in severe viral pneumonia. Molecular tools significantly increase detection of respiratory pathogens^7-10^ and have been used to safely discontinue antibiotics in patients with severe CAP requiring mechanical ventilation.^10^ Samples collected by nasopharyngeal swab or endotracheal aspiration are inferior to sampling of the alveolar space by bronchoalveolar lavage (BAL) for the detection of respiratory pathogens.^11-14^ However, concerns regarding the operator-safety of performing BAL in patients with COVID-19 pneumonia have precluded its use in many centers.^15^ Hence, the prevalence of initial bacterial superinfection and subsequent ventilator-associated pneumonia (VAP) in patients with SARS-CoV-2 pneumonia are not known.^16,17^ Moreover, the spectrum and antibiotic susceptibility of superinfecting pathogens remains undefined. Patients may therefore be exposed to unnecessary empirical broad-spectrum antibiotics, use of which has been associated with late infection with resistant organisms.^16^

We modified the standard bronchoscopic BAL technique in patients intubated for COVID-19 pneumonia-related respiratory failure to minimize operator exposure to infectious aerosols.^18^ We retrospectively analyzed data from a cohort managed during the initial local COVID-19 surge to determine the prevalence and microbiology of bacterial superinfection at the time of intubation and subsequent bacterial VAP.

## Methods

### Patient cohort

We conducted this retrospective, observational study at Northwestern Memorial Hospital (NMH), a quaternary, acute care hospital. Patients discharged from the hospital between March 1 and June 30, 2020 were included in the cohort. Patients who were SARS-CoV-2-positive but intubated for reasons other than pneumonia (surgical procedures, intoxication, etc.) were excluded by formal adjudication of at least two critical care physicians.

A BAL early after endotracheal intubation is our center’s standard of clinical care for patients with suspected pneumonia. We modified our standard bronchoscopic BAL technique for ventilated patients in order to minimize aerosol generation.^18^ We defined early bacterial superinfection as detection by culture or multiplex PCR of a respiratory pathogen known to cause pneumonia, in addition to SARS-CoV-2, on a BAL specimen collected within 48 hours after intubation. We defined VAP as detection of a new respiratory pathogen in BAL fluid obtained more than 48 hours after intubation.

### Endpoints

We examined two primary endpoints: 1) the prevalence of bacterial superinfection at the time of intubation and 2) the incidence rate of subsequent VAP over the duration of mechanical ventilation. Secondary endpoints included the emergence of pathogens demonstrating resistance to antimicrobial therapies, clinical outcomes based on infection status, and the use of antibiotics.

Beta-lactam antibiotics are considered the backbone of pneumonia treatment.^19^ Multidrug resistance (MDR) was defined by the need for a carbapenem for Gram-negative pathogens or vancomycin/linezolid for *S. aureus*. For each day of mechanical ventilation, we measured the spectrum of antibacterial antibiotic therapy using a Narrow Antibiotic Treatment (NAT) score developed for CAP treatment.^20^ Briefly, standard CAP treatment of ceftriaxone and azithromycin was assigned a score of 0, monotherapy with either was assigned a −1 score, and no antibiotic therapy was assigned a −2 score; broader spectrum antibiotics were assigned progressively higher positive scores (see Supplementary material).

### Statistical Analysis

Data were analyzed using custom scripts in R 4.0.2 using tidyverse 1.3.1 (The R Foundation for Statistical Computing, http://www.R-project.org). All plotting was performed using ggplot2 3.3.2. Cohort characteristics are reported as median and interquartile range (IQR) for quantitative variables and percentages for categorical variables. Median NAT scores were compared using non-parametric methods (Kruskal-Wallis, Wilcoxon Rank-Sum test, and Fisher’s Exact test as appropriate). In cases of multiple testing, P-values were corrected using false-discovery rate (FDR) correction. Adjusted P-values < 0.05 were considered significant. Two-sided statistical tests were performed in all applicable cases.

## Results

### Clinical features of the cohort

From March 1 to June 30, 2020, we cared for 196 patients intubated for severe SARS-CoV-2 pneumonia; the 179 discharged from the hospital by June 30 were included in the analysis. Characteristics and outcomes of these patients are demonstrated in Table 1. Patients transferred from an external hospital constituted 28% of the population and were more likely to be managed with ECMO, had higher mortality, and a shorter duration of mechanical ventilation.

**Table 1.** Demographics, clinical characteristics, and outcomes by early BAL status.

|  | Early BAL |  |  |  |  |
| --- | --- | --- | --- | --- | --- |
|  | Total | Without Superinfection | With Superinfection | No Early BAL | P-value |
| **Number** | 179 | 105 | 28 | 46 | - |
| **Age (median, IQR)** | 62.4 (22.5) | 62.1 (21.9) | 63.8 (19.9) | 59.9 (22.0) | 0.96 |
| **Male** | 110 (61.5%) | 69 (65.7%) | 22 (78.6%) | 19 (41.3%) | 0.003 |
| **Race/Ethnicity** |  |  |  |  | 0.74 |
| **White** | 37 (20.7%) | 25 (23.8%) | 5 (17.9%) | 7 (15.2%) |  |
| **Hispanic** | 63 (35.2%) | 38 (36.2%) | 8 (28.6%) | 17 (37.0%) |  |
| **Black** | 60 (33.5%) | 31 (29.5%) | 13 (46.4%) | 16 (34.8%) |  |
| **Asian** | 8 (4.5%) | 5 (4.8%) | 0(0.00%) | 3 (6.5%) |  |
| **Other** | 11 (6.2%) | 6 (5.7%) | 2 (7.14%) | 3 (6.5%) |  |
| **External Transfers** | 33 (18.4%) | 11 (10.5%) | 5 (17.9%) | 17 (37.0%) | 0.001 |
| **Admission BMI** |  |  |  |  | 0.22 |
| **<24.9** | 28 (15.6%) | 14 (13.3%) | 6 (21.4%) | 8 (17.4%) |  |
| **25 – 29.9** | 48 (26.8%) | 29 (27.6%) | 7 (25.0%) | 12 (26.1%) |  |
| **30 – 39.9** | 71 (39.7%) | 47 (44.8%) | 6 (21.4%) | 18 (39.1%) |  |
| **> 40** | 32 (17.9%) | 15 (14.3%) | 9 (32.1%) | 8 (17.4%) |  |
| **SOFA Score - median, IQR** | 7 (4-9) | 7 (3-9) | 8 (3.75-10) | 7.5 (6-9) | 0.43 |
| **Comorbidities** |  |  |  |  |  |
| **Diabetes** | 80 (44.7%) | 49 (46.7%) | 13 (46.4%) | 18 (39.1%) | 0.71 |
| **Hypertension** | 105 (58.7%) | 65 (61.9%) | 17 (60.7%) | 23 (50.0%) | 0.42 |
| **Atrial fibrillation** | 16 (8.9%) | 12 (11.4%) | 3 (10.7%) | 1 (2.2%) | 0.15 |
| **Coronary artery disease** | 21 (11.7%) | 10 (9.5%) | 6 (21.4%) | 5 (10.9%) | 0.22 |
| **Heart failure** | 24 (13.4%) | 13 (12.4%) | 3 (10.7%) | 8 (17.4%) | 0.68 |
| **COPD** | 16 (8.8%) | 10 (9.5%) | 2 (7.1%) | 4 (8.7%) | 1 |
| **Asthma** | 11 (6.2%) | 3 (2.9%) | 2 (7.1%) | 6 (13.0%) | 0.044 |
| **Obstructive sleep apnea** | 26 (14.5%) | 16 (15.2%) | 5 (17.9%) | 5 (10.9%) | 0.66 |
| **Solid organ transplant** | 11 (6.2%) | 10 (9.5%) | 0 (0.0%) | 1 (2.2%) | 0.11 |
| **BMT/Heme malignancy** | 3 (1.7%) | 3 (2.9%) | 0 (0.0%) | 0 (0.0%) | 0.73 |
| **Other cancer** | 15 (8.4%) | 10 (9.5%) | 2 (7.1%) | 3 (6.5%) | 0.93 |
| **Chronic hemodialysis** | 14 (7.8%) | 12 (11.4%) | 1 (3.6%) | 1 (2.2%) | 0.12 |
| **Chronic kidney disease** | 30 (16.8%) | 22 (21.0%) | 4 (14.3%) | 4 (8.7%) | 0.16 |
| **PE/DVT** | 12 (6.7%) | 7 (6.7%) | 1 (3.6%) | 4 (8.7%) | 0.77 |
| **CVA** | 8 (4.5%) | 5 (4.8%) | 1 (3.6%) | 2 (4.3%) | 1 |
| **Cirrhosis** | 3 (1.7%) | 2 (1.9%) | 0 (0.0%) | 1 (2.2%) | 1 |
| **Active smoker** | 4 (2.4%) | 3 (3.0%) | 1 (3.6%) | 0 (0.0%) | 0.63 |
| **Charleson Comorbidity Index**<br>**Median (median, IQR)** | 2 (4) | 2 (5) | 1(2.25) | 1.5(2.75) | 0.66 |
| **Biomarkers <sup>1</sup>** |  |  |  |  |  |
| **C-Reactive Protein** | 16.9 (5.7) | 17.60 (13.9) | 19.4 (11.7) | 14.3 (15.3) | 0.077 |
| **Procalcitonin** | 0.43 (1.83) | 0.43 (1.63) | 0.54 (1.92) | 0.40 (2.49) | 0.98 |
| **WBC** | 8.60 (5.65) | 9.20 (5.70) | 10.05 (5.70) | 7.60 (6.0) | 0.16 |
| **Absolute lymphocytes** | 0.90 (.60) | 0.90 (0.73) | 1.10 (0.70) | 0.95 (.40) | 0.74 |
| **Absolute neutrophils** | 7.60 (5.7) | 7.50 (5.17) | 10.00 (6.45) | 6.50 (5.28) | 0.027 |
| **D-dimer** | 601 (820) | 590 (649) | 542 (1474) | 719 (1999) | 0.65 |
| **Ferritin** | 8556 (1235) | 902 (1482) | 1110 (867) | 718 (850) | 0.36 |
| **Troponin** | 0.03 (0.06) | 0.03 (0.05) | 0.03 (0.07) | 0.03 (0.05) | 0.73 |
| **Management** |  |  |  |  |  |
| **Proned** | 89 (49.7%) | 51 (48.6%) | 14 (50.0%) | 24 (52.2) | 0.93 |
| **ECMO** | 17 (9.5%) | 7 (6.7%) | 2 (7.1%) | 8 (17.4) | 0.13 |
| **Anti-IL6r study<sup>2</sup>** | 17 (9.5%) | 12 (11.4%) | 2 (7.1%) | 3 (6.5) | 0.72 |
| **Anti-IL6r off-label** | 30 (16.8%) | 20 (19.0%) | 2 (7.1%) | 8 (17.4) | 0.40 |
| **Remdesivir study<sup>3</sup>** | 15 (8.4%) | 7 (6.7%) | 3 (10.7%) | 5 (10.9) | 0.56 |
| **Remdesivir EUA** | 18 (10.1%) | 10 (9.5%) | 1 (3.6%) | 7 (15.2) | 0.30 |
| **HCQ** | 40 (22.1%) | 20 (19.0%) | 5 (17.9%) | 15 (32.6) | 0.16 |
| **Corticosteroids<sup>4</sup>** | 58 (32.0%) | 29 (27.6%) | 11 (39.3%) | 18 (39.1) | 0.26 |
| **Hospital Course** |  |  |  |  | 0.068 |
| **Ventilation Duration, days<sup>5</sup>** | 13.0 (18.5) | 13.0 (18.7) | 16.7 (22.3) | 13.2 (12.6) | 0.11 |
| **ICU LOS, days<sup>6</sup>** | 16.0 (18.0%) | 16.9 (18.1%) | 17.3 (21.6%) | 13.7 (14.2%) | 0.13 |
| **Hospital LOS, days** | 25.7 (19.0%) | 26.9 (18.8%) | 30.2 (17.7%) | 21.1 (17.6%) | 0.03<sup>7</sup> |
| **VAP** | 72 (40.2%) | 38 (36.2%) | 14 (50.0%) | 20 (43.5%) | 0.37 |
| **- MDR pathogen** | 19 (10.6%) | 9 (8.6%) | 6 (21.4%) | 4 (8.7%) | 0.15 |
| **Tracheostomy** | 48 (26.8%) | 27 (25.7%) | 11 (39.3%) | 10 (21.7%) | 0.25 |
| **Chronic respiratory support on discharge<sup>8</sup>** | 20 (11.2%) | 11 (10.5%) | 6 (21.4%) | 3 (6.5%) | 0.15 |
| **Renal Replacement Therapy during admission** | 49 (27.4%) | 32 (30.5%) | 9 (32.1%) | 8 (17.4%) | 0.21 |
| **New chronic HD on discharge** | 5 (2.8%) | 3 (2.9%) | 2 (7.1%) | 0 (0.0%) | 0.16 |
| **Discharge Outcomes** |  |  |  |  |  |
| **Death** | 34 (19.0%) | 17 (16.2%) | 3 (10.7%) | 14 (30.4%) |  |
| **LTACH** | 25 (14.0%) | 13 (12.2%) | 9 (32.1%) | 3 (6.5%) |  |
| **Nursing home (SNF)** | 11 (6.2%) | 8 (7.6%) | 0 (0.0%) | 3 (6.5%) |  |
| **Acute inpatient rehab** | 25 (14.0%) | 14 (13.3%) | 4 (14.3%) | 7 (15.2%) |  |
| **Home** | 84 (46.0%) | 52 (50.5%) | 12 (42.9%) | 19 (41.3%) |  |

<sup>1</sup> Biomarkers at intubation (or date of transfer to NMH for cases of external transfers); <sup>2</sup>randomized ranged from 4:1 to 2:1,
<sup>3</sup>randomized 1:1; <sup>4</sup>specifically for COVID; <sup>5</sup>As total time under ventilation, including time in other institutions before transfer, Median and IQR; <sup>6</sup>Length of stay at Northwestern Memorial Hospital system; for external transfers, when exact ICU length of stay was not known, date of intubation was used for start of ICU stay; <sup>7</sup>Northwestern Memorial Hospital system only, not meaningful comparison;
<sup>8</sup>Long Term Acute Care Hospital (LTACH) on ventilator, lung transplant, home ventilation

### Prevalence of superinfecting pathogens

The majority (74.3%, 133/179) of patients had a BAL obtained within the initial 48 hours of intubation. External transfer patients were less likely to have an early BAL (51.5% vs. 74.3%, *p*<.001). The median duration of ventilation among external transfer patients at the time of transfer was two days, and therefore many were outside the window for an early BAL by our definition. No differences in other baseline characteristics of the population that did not undergo early BAL were found.

Of patients who underwent an early BAL, 21.1% (28/133) had a documented bacterial superinfection. Of early BAL patients, 37 (27.8%) were admitted to the hospital for >48 hours, meeting the definition of suspected hospital-acquired pneumonia (HAP). Despite enrichment of the cohort for HAP, etiologies of early post-intubation pneumonia were typical of CAP, with *Streptococcus* species and methicillin-susceptible *S. aureus* (MSSA) combined accounting for 79% (22/28) of cases. Polymicrobial infections were common with 51 pathogens detected in 28 BAL fluid samples. Only three patients had pathogens resistant to standard CAP antibiotics – one *Stenotrophomonas maltophilia* and two MRSA. Pneumocystis was detected in one HIV patient well-controlled on antiretroviral treatment.

Standard clinical parameters (e.g., fever) or blood biomarkers did not distinguish between patients with and without early bacterial superinfection (Table 2). The cellular composition of the BAL fluid in patients with SARS-CoV-2 pneumonia differed compared to historic patients in our center with pneumonia caused by other respiratory pathogens, with 55.6% having >10% lymphocytes (Supplemental Figure 1).^21^ Nevertheless, cellular composition of BAL fluid alone was insufficient to distinguish patients with superinfection from those without, as BAL neutrophils exceeded 50% in 36.5% of cases without bacterial superinfection, and lymphocytes exceeded 10% in half of cases with or without bacterial superinfection.

**Table 2.** Early BAL characteristics and pathogens.

|  | With<br>Superinfection<br>N=28 | Without<br>Superinfection<br>N=105 | p-value |
| --- | --- | --- | --- |
| T max prior to BAL | 102.0 (2.8) | 100.6 (2.9) | 0.39 |
| White blood count | 9.4 (6.0) | 8.8 (6.0) | 0.66 |
| Absolute neutrophil count | 8.3 (6.3) | 7.1 (5.3) | 0.39 |
| Absolute lymphocyte count | 0.9 (0.7) | 0.9 (0.7) | 0.66 |
| NLR (neutrophil to lymphocyte ratio)<sup>1</sup> | 8 (7.5) | 7.1 (5.8) | 0.72 |
| C-reactive protein<sup>2</sup> | 16.9 (13.1) | 17.7 (14.1) | 1.00 |
| D-dimer<sup>3</sup> | 630 (1507) | 550 (604) | 1.00 |
| Ferritin | 1030 (1298) | 726 (1331) | 0.95 |
| Antibiotics > 24 hours before BAL (%)<sup>4</sup> | 5 (18%) | 38 (36%) | 0.57 |
| Hospitalization > 48 hours (%) | 4 (14%) | 33 (31%) | 0.47 |
| BAL characteristics |  |  |  |
| RBC (/mm<sup>3</sup>)<sup>5</sup> | 27078 (3715) | 2375 (6275) | 1.00 |
| WBC (/mm<sup>3</sup>)<sup>6</sup> | 258 (826) | 164 (253) | 0.39 |
| Neutrophils (%)<sup>7</sup> | 42 (55%) | 41 (43%) | 0.72 |
| Neutrophil % > 50<sup>7</sup> | 10 (36%) | 36 (34%) | 1.00 |
| Lymphocytes (%)<sup>7</sup> | 11 (20%) | 14 (19%) | 0.66 |
| Lymphocyte % > 10<sup>7</sup> | 14 (50%) | 60 (57%) | 0.72 |
| Macrophages (%)<sup>7</sup> | 7 (18%) | 14 (26%) | 0.39 |
| Monocytes (%)<sup>7</sup> | 8 (15%) | 9 (12%) | 0.66 |
| Others (%)<sup>7,9</sup> | 3 (8%) | 3 (7%) | 0.72 |
| Amylase (IU/L)<sup>8</sup> | 29 (46) | 16 (32) | 0.47 |
| Amylase > 105 IU/L (%)<sup>8</sup> | 2 (7%) | 2 (2%) | 0.66 |
| Microbiology results N (%) |  |  |  |
| *Staphylococcus aureus* -sensitive | 11 (39%) |  |  |
| -resistant | 2 (7%) |  |  |
| *Viridans streptococcus* | 10 (36%) |  |  |
| *Streptococcus agalactiae* | 3 (11%) |  |  |
| *Streptococcus pneumoniae* | 3 (11%) |  |  |
| Other *Streptococcus* species | 2 (7%) |  |  |
| *Haemophilus influenzae* | 2 (7%) |  |  |
| *Stomatococcus* species | 2 (7%) |  |  |
| *Enterococcus* species | 1 (4%) |  |  |
| *Klebsiella oxytoca* | 1 (4%) |  |  |
| *Moraxella catarrhalis* | 1 (4%) |  |  |
| *Proteus mirabilis* | 1 (4%) |  |  |
| *Serratia marcescens* | 1 (4%) |  |  |
| *Stenotrophomonas maltophilia* | 1 (4%) |  |  |

Data unavailable for the following number of patients: <sup>1</sup> No Superinfection (NS) group=4;
<sup>2</sup>Superinfection group (S) =2, NS=1; <sup>3</sup> NS=2; <sup>4</sup> S=3; <sup>5</sup> S=2. NS=18; <sup>6</sup> S=1, NS=7; <sup>7</sup>S=1, NS=6;
<sup>8</sup>S=16, NS=58. <sup>9</sup>plasma cells, eosinophils, any other.

**Figure 1.**
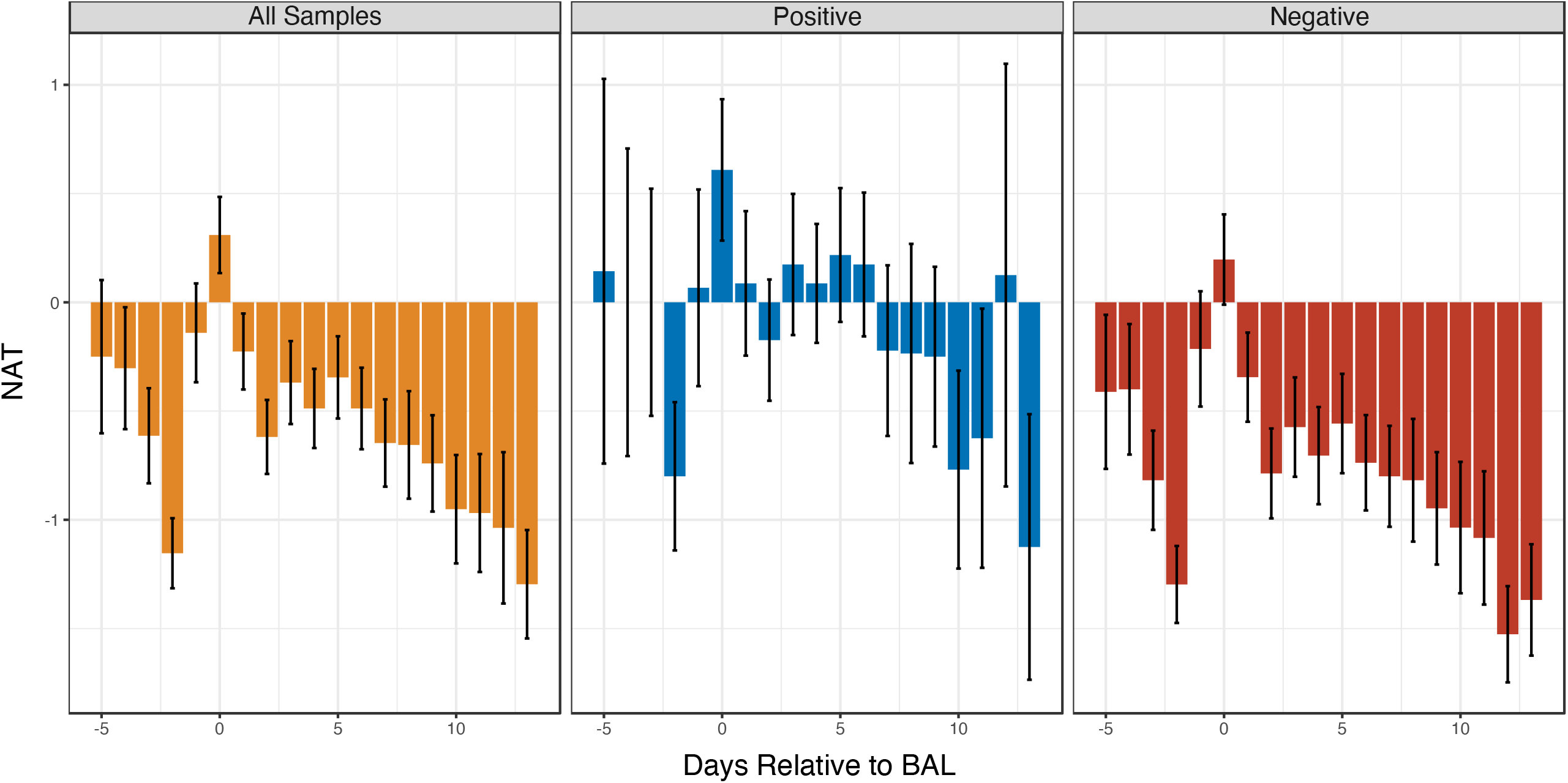
Median Narrow Antibiotic Therapy (NAT) score in response to BAL results overall, and in response to positive and negative BAL results for patients undergoing bronchoscopy within 48 hours of intubation.

Median NAT scores demonstrate that BAL results were associated with changes in antibiotic management (Figure 1). Among all early BAL patients, the median daily NAT score in the first 7 days was −1 (95% CI –1.5, −0.5), significantly different than guideline-recommended treatment (NAT >0, *p*<.001). While the median daily NAT score for patients with a positive BAL (median – 1, 95%CI –1, 0) was not different than guideline-recommended treatment (Figure 1a), the median daily NAT score for patients with a negative BAL was significantly lower (median –1.5, 95%CI –1.5, −0.5; *p*<.001, Figure 1b). The median difference between NAT scores for patients with positive and negative BAL results was statistically significant (median difference −1, 95%CI −1, 0; *p*=.001).

### Episodes of ventilator-associated pneumonia

An additional 246 BALs were performed on the 162 patients that remained intubated for more than 48 hours (Figure 2a). Only 18 (11.1%) patients never underwent a BAL after 48 hours of intubation. Patients who did not have a BAL had a lower median duration of ventilation (5.0 days, IQR 3.0 to 8.5) than those who did (14.0 days, IQR 8 to 27.0; *p*<.001).

**Figure 2.**
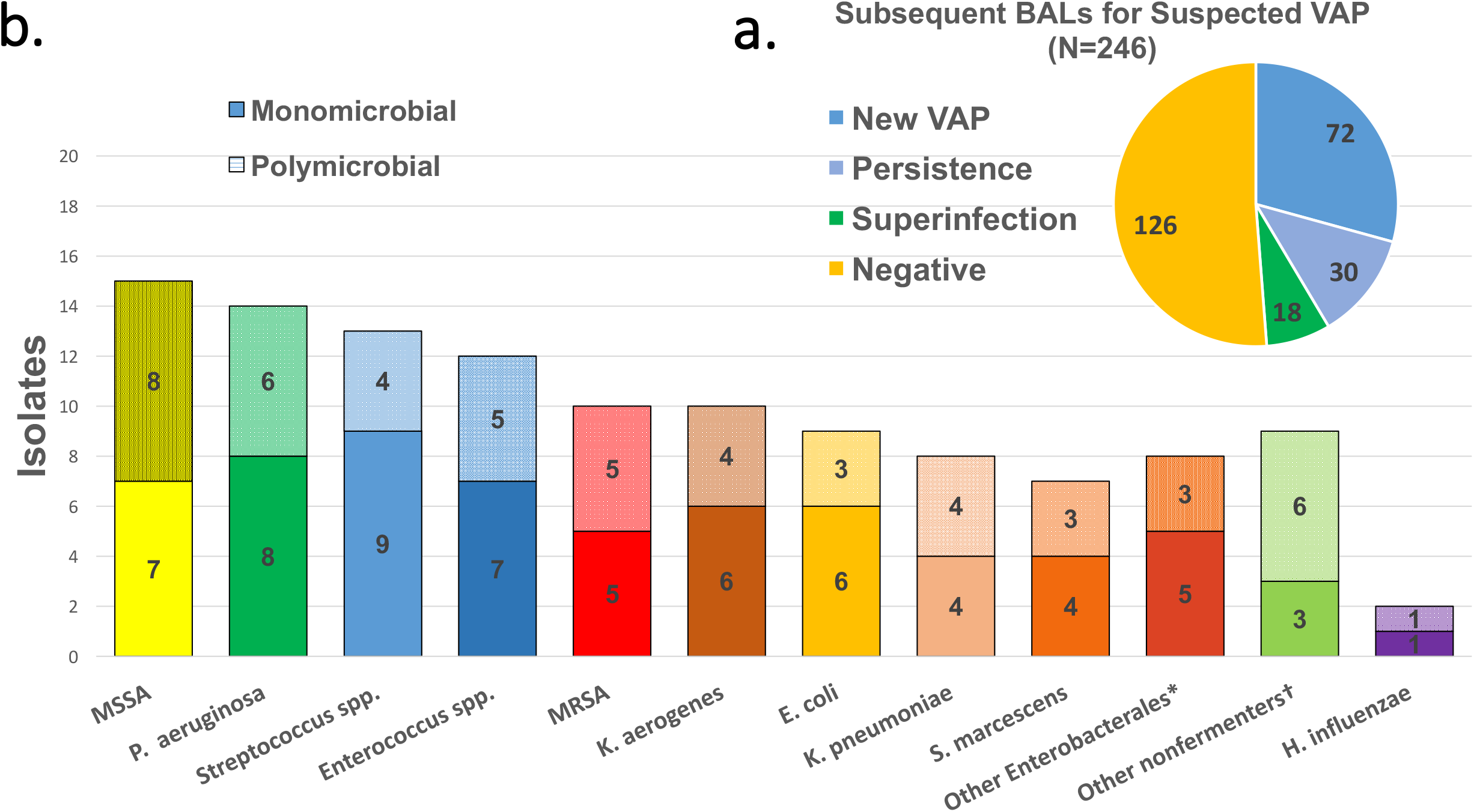
BAL results for suspected VAP (a) and pathogens detected in positive BALs (b). Solid bars are pathogens detected in monomicrobial episodes while crosshatched are presence in polymicrobial pneumonias.

VAP was diagnosed in 120 BALs from 72 unique patients (44.4% of all patients) while 126 (51.2%) BALs had no evidence of VAP. The first episode of VAP occurred an average of 10.8 days post-intubation. Of these patients, 20.8% (15/72) developed a second VAP a median of 9.7 days after the first episode; three patients developed a third. Persistence of a previously identified pathogen causing VAP was found in 30 additional BALs obtained 1-34 days (median 10.7) after a previous BAL. Patients with a documented early bacterial superinfection had more VAPs and more VAP secondary to MDR pathogens, although these differences were not significant (Table 1).

Clinical characteristics and blood biomarkers among patients with VAP did not differ from those with suspected VAP but a negative BAL. In contrast, the cellular composition of the BAL fluid showed a significantly higher percentage of neutrophils and lower percentage of lymphocytes in patients with VAP compared to those without (Table 3).

**Table 3.** Late BAL characteristics (positive vs. negative)

| **Table 3** | **With VAP** | **Without VAP** | **p-value** |
| --- | --- | --- | --- |
| **Lab/vitals at time of BAL** | **n = 120** | **n = 126** |  |
| T max prior to BAL | 100.8 (2.7) | 100.6 (1.9) | 0.97 |
| WBC | 12.8 (8.4) | 11.8 (6.6) | 0.45 |
| Absolute neutrophil count | 8.7 (8.5) | 8.4 (8.2) | 0.55 |
| Absolute lymphocyte count | 1.0 (0.9) | 1.0 (1.0) | 0.83 |
| NLR (neutrophil to lymphocyte ratio)<sup>1</sup> | 7.6 (12.6) | 8.8 (7.4) | 0.97 |
| CRP<sup>2</sup> | 14.8 (16.7) | 15.3 (21.5) | 0.51 |
| D-dimer<sup>3</sup> | 2356 (2699) | 2091 (2276) | 0.61 |
| Ferritin | 559 (1013) | 586 (1127) | 0.95 |
| Antibiotics > 24 hours before BAL (%)<sup>4</sup> | 32 (27%) | 29 (23%) | 0.83 |
| Day of hospitalization for BAL | 16 (11) | 10 (11) | <.0003 |
| **BAL characteristics** |  |  |  |
| RBC (/mm<sup>3</sup>)<sup>5</sup> | 2525 (8390) | 1450 (4475) | 0.33 |
| WBC (/mm<sup>3</sup>)<sup>6</sup> | 488 (1635) | 242 (440) | 0.002 |
| Neutrophils (%)<sup>7</sup> | 77 (41%) | 48 (48%) | <.0001 |
| Neutrophil % > 50 %<sup>7</sup> | 83 (69%) | 57 (45%) | <.0004 |
| Lymphocytes (%)<sup>7</sup> | 5 (15%) | 14 (24%) | <.0003 |
| Lymphocyte % > 10 %<sup>7</sup> | 38 (32%) | 75 (59%) | <.0003 |
| Macrophages (%)<sup>7</sup> | 6 (13%) | 12 (23%) | 0.012 |
| Monocytes (%)<sup>7</sup> | 3 (5%) | 5 (8%) | 0.13 |
| Others (%)<sup>7,9</sup> | 1 (4%) | 3 (6%) | 0.002 |
| Amylase<sup>8</sup> | 31 (76) | 20 (75) | 0.45 |
| Number (%) Amylase > 105 IU/L<sup>8</sup> | 15 (13%) | 8 (6%) | 0.32 |

Data unavailable at the time of BAL for the following number of patients: <sup>1</sup>S=16, NS=19; <sup>2</sup>S=4, NS=6; <sup>3</sup>S=32, NS=36; <sup>4</sup>S=5, NS=2; <sup>5</sup>S=11, NS=13; <sup>6</sup>S=9, NS=5; <sup>7</sup>S=5, NS=2; <sup>8</sup>S=60, NS=65. <sup>9</sup>plasma cells, eosinophils, any other.

Figure 2b illustrates the most common pathogens causing VAP in the cohort. Monomicrobial VAP was more common for the first VAP episode (56/72, 77.8%) than for subsequent episodes (8/18, 44.4%, *p*=.005). Figure 3a shows the cumulative contribution of different organisms to the development of VAP over the duration of mechanical ventilation. Only 15 of the 72 (20.8%) initial VAP etiologies were MDR pathogens, including nine Gram-negative pathogens resistant to piperacillin/tazobactam and cefepime and six episodes attributable to MRSA. A substantial number of Gram-negative VAPs (48.6%) could be treated with cefazolin or ceftriaxone monotherapy. The 18 superinfections were only slightly more resistant with 33% MDR pathogens - two MRSA and four Gram-negative pathogens resistant to piperacillin/tazobactam and cefepime.

**Figure 3a.**
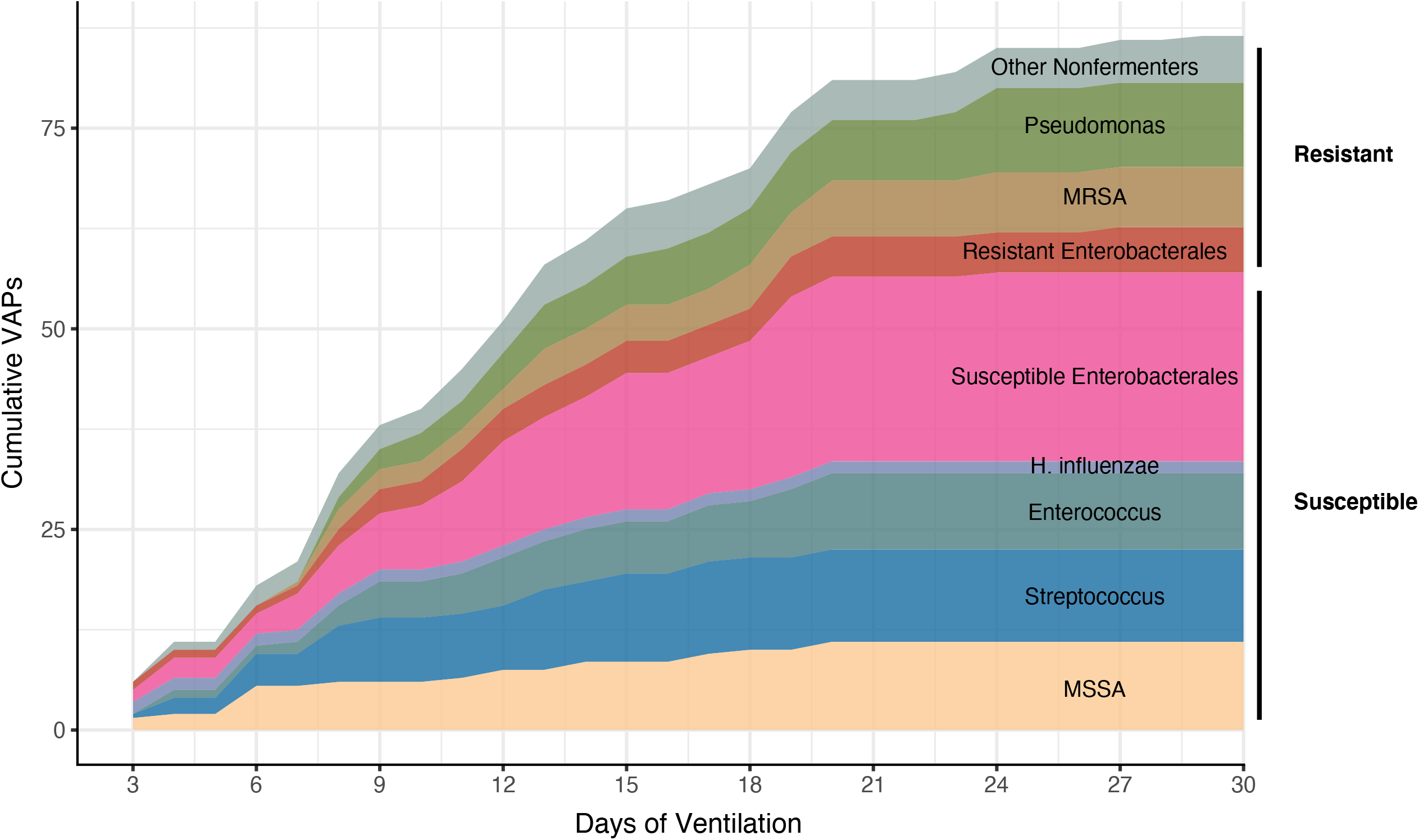
Cumulative VAPs by etiology and resistance pattern. For Enterobacterales, resistant isolates required carbapenem or broader spectrum beta-lactam treatment. MRSA- methicillin-resistant *S. aureus*, MSSA-methicillin-susceptible *S. aureus*

**Figure 3b.**
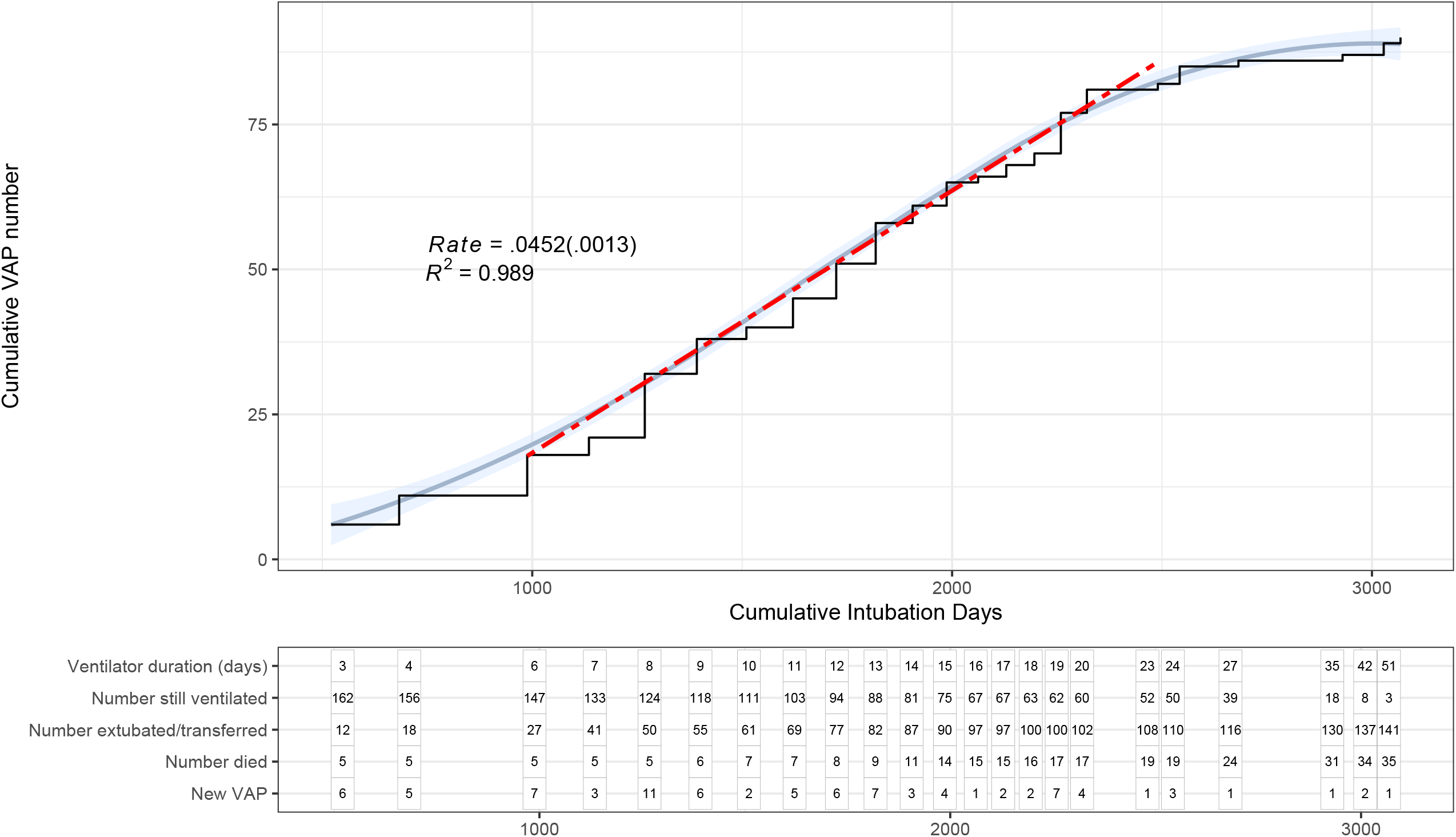
Incidence of VAP. Cumulative new VAP diagnoses per cumulative ventilator days. Individual patients can have more than one VAP episode.

The overall VAP incidence rate in this cohort was 45.2 episodes/1000 days of mechanical ventilation. The VAP incidence rate was linear over cumulative days on mechanical ventilation until the number of patients still on mechanical ventilation became very low (Figure 3b). The distribution of ventilator day of VAP onset is shown in Supplemental Figure 2.

### Clinical Outcomes

Overall hospital mortality was 19%. Mortality of external transfers was greater than patients managed wholly at NMH (34.3% vs. 15.3% respectively, OR 2.87 [95% CI 1.13, 7.11], *p*=.01). Mortality in patients with documented bacterial superinfection at the time of intubation was not higher than in those with only SARS-CoV-2 detected (Table 1). However, early bacterial superinfection was associated with more prolonged ventilation and corresponding tracheostomy and chronic respiratory support.

## Discussion

Using sensitive and specific testing of alveolar samples, we defined the microbial epidemiology of bacterial superinfection in patients with severe SARS-CoV-2 pneumonia requiring mechanical ventilation. Current guidelines recommend empirical administration of antibiotics to all patients with severe SARS-CoV-2 pneumonia.^3,11,22^ As the prevalence of bacterial superinfection was 21%, guideline-based antibiotic management would have resulted in substantial overtreatment. Despite the high subsequent VAP incidence rate of 45.2/1000 days of ventilation, the majority of VAP pathogens in our cohort were not MDR pathogens. As a result, BAL-based antibiotic management resulted in less frequent and more narrow-spectrum antibiotic therapy compared with current guidelines for the empirical treatment of pneumonia.

Because time from onset of symptoms to need for mechanical ventilation is longer in patients with SARS-CoV-2 pneumonia compared with other severe CAPs, many patients meet the definition for HAP at the time of intubation (27.8% of our cohort). Despite this, we found predominantly bacterial isolates characteristic of CAP, predominantly *Streptococcus* species and MSSA. These species remained prevalent even in early VAP cases (days 3-7 of intubation). The low prevalence and CAP-like spectrum of bacterial superinfections suggests that most patients intubated with SARS-CoV-2 pneumonia do not require antibiotics and those that do, can often be managed with narrow-spectrum therapy. The relatively low prevalence of bacterial superinfection may partially explain why early antibiotics are reported to have no impact on COVID-19 pneumonia mortality.^23^

Use of sensitive and specific PCR and culture techniques to analyze BAL fluid likely contributed to the higher prevalence (21%) of bacterial superinfection at the time of intubation in our study compared with others. A recent meta-analysis, in which the diagnosis of pneumonia relied on cultures of endotracheal aspirates or sputum, reported the prevalence of bacterial superinfection was 14% in severe SARS-CoV-2 pneumonia.^6^ Other small BAL-based prevalence studies found an 8-10% rate of early bacterial superinfection.^24,25^ In a small cohort of COVID-19 patients, investigators used multiplex PCR to analyze sputum and endotracheal aspirates, reporting co-infections in 40.6%.^26^ However, concern for false positives has been raised for these types of samples.^27^ Our combination of a true alveolar sample combined with sophisticated laboratory testing algorithms likely provides a more accurate estimate of prevalence.

Few studies have addressed VAP complicating severe SARS-CoV-2 pneumonia, with prevalence estimates ranging between 28.6% and 38%.^25,28-30^ In our cohort, 44% were diagnosed with at least one episode of VAP. The slightly higher prevalence rate of VAP is likely attributable to the use of the multiplex PCR technology.^31^

We are aware of no other reports of the incidence of VAP in severe SARS-CoV-2 pneumonia. The VAP incidence rate of 45.2/1000 ventilator days in our SARS-CoV-2 pneumonia cohort is substantially higher than that reported in other critically ill patients receiving mechanical ventilation.^32,33^ In addition to enhanced diagnosis with use of multiplex PCR technology, this high VAP incidence may reflect clinical features unique to SARS-CoV-2 pneumonia.

Specifically, as the incidence of VAP is constant over the duration of ventilation in our cohort, the relatively long length of stay in patients with SARS-CoV-2 pneumonia combined with their relatively low mortality (a competing outcome for VAP) would increase both incidence and prevalence of VAP. Despite the relatively high rates of VAP, only 11% of organisms were classified as MDR, less than reported in most VAP studies.

The clinical rationale supporting guideline-based recommendations for empirical antibiotics in patients with severe viral pneumonia includes concern that failure to treat bacterial superinfection could increase mortality, and the hypothesis that early antibiotic therapy might prevent subsequent bacterial superinfection pneumonia. In our cohort of severe SARS-CoV-2 pneumonia patients, bacterial superinfection at the time of intubation was not associated with increased mortality, contrasting with reports in patients with severe influenza^34^ and other small cohorts of patients with SARS-CoV-2 pneumonia.^28^ However, bacterial superinfections were associated with a longer duration of mechanical ventilation, with its associated complications including VAP and tracheostomy.

Any clinical benefit of greater BAL diagnostic accuracy is dependent on clinical confidence in the results sufficient to not initiate, narrow or discontinue empirical antibiotic therapy; ignoring results (continuing empirical antibiotic management) will not result in benefit.^35,36^ After BAL, clinicians in our center narrowed or discontinued antibiotic therapy in most patients, suggesting they were comfortable using these data for antibiotic decisions. For example, almost 50% of first VAP patients were managed with cefazolin or ceftriaxone monotherapy. Despite less use of antibiotics compared to guideline-based care, the overall low mortality in our cohort suggests important bacterial infections were not missed. These clinical decisions are unlikely attributable to clinical features unrelated to the BAL procedure as SARS-CoV-2 infection alone is associated with prolonged fever, radiographic infiltrates and elevated inflammatory markers commonly used in diagnostic algorithms for pneumonia.

## Limitations

Like all single-center studies, the generalizability of our findings to other centers is unclear. While BAL cultures are more specific than endotracheal aspirates,^12,13^ sensitivity may be adversely affected by prior antibiotic therapy; our data may still underestimate the true frequency of early superinfection. Other aspects of COVID-19 pneumonia management, such as timing of intubation, use of noninvasive ventilation, ventilator strategy, and availability of ECMO, may affect the rate of bacterial pneumonia and may therefore affect pneumonia rates. Our relatively low mortality despite a longer length of stay compared to that reported from other centers will tend to increase the observed prevalence of subsequent VAP.

## Summary

Superinfection bacterial pneumonia was present at the time of intubation in 21% of severe SARS-CoV-2 pneumonia cases. The prevalence of subsequent VAP was 44% with an incidence rate of 45.2/1000 ventilator days. Superinfection bacterial pneumonias at time of intubation and early VAPs were predominantly caused by pathogens usually associated with CAP and susceptible to narrow-spectrum antibiotic therapy. Empirical treatment of severe SARS-CoV-2 pneumonia based on current guideline-based recommendations would have resulted in substantial antibiotic overuse in our cohort.

## Data Availability

Data are available upon reasonable request.

## Acknowledgements

The Northwestern University COVID Investigators include:

A. Christine Argento, Ajay A. Wagh, Alexander V. Misharin, Alexandra C. McQuattie-Pimentel, Alexis Rose Wolfe, Alvaro Donayre, Ankit Bharat, Anna E. Pawlowski, Anne R. Levenson, Anthony M. Joudi, Benjamin D. Singer, Betty Tran, Catherine A. Gao, Chao Qi, Chiagozie O. Pickens, Chitaru Kurihara, Clara J Schroedl, Daniel Meza, Daniel Schneider, David A. Kidd, David D. Odell, David W. Kamp, Elizabeth S. Malsin, Emily M. Leibenguth, Eric P. Cantey, Gabrielle Y. Liu, GR Scott Budinger, Helen K. Donnelly, Isaac A. Goldberg, Jacob I. Sznajder, Jacqueline M. Kruser, James M. Walter, Jane E. Dematte, Jason M. Arnold, John Coleman, Joseph I. Bailey, Joseph S. Deters, Justin A. Fiala, Katharine Secunda, Kaitlyn Vitale, Khalilah L. Gates, Kristy Todd, Lindsey D. Gradone, Lindsey N.Textor, Lisa F. Wolfe, Lorenzo L. Pesce, Luisa Morales-Nebreda, Madeline L. Rosenbaum, Manu Jain, Marc A. Sala, Mary Carns, Marysa V. Leya, Mengjia Kang, Michael J. Alexander, Michael J. Cuttica, Michelle Hinsch Prickett, Natalie Jensema, Nicole Borkowski, Nikolay S. Markov, Orlyn R. Rivas, Paul A. Reyfman, Peter H. S. Sporn, Prasanth Nannapaneni, Rachel B. Kadar, Rachel M. Kaplan, Rade Tomic, Radhika Patel, Rafael Garza-Castillon, Ravi Kalhan, Richard G. Wunderink, Rogan A. Grant, Romy Lawrence, Ruben J. Mylvaganam, Samuel S. Kim, Sanket Thakkar, Sean B. Smith, SeungHye Han, Sharon R. Rosenberg, Susan R. Russell, Sydney M. Hyder, Taylor A. Poor, Theresa A. Lombardo, Zasu M. Klug. ORCHID identifiers and affiliations are found in Supplementary information.

