## Supplementary for "Bacterial superinfection pneumonia in SARS-CoV-2 respiratory failure"

Table of Contents:

1. Table 1. NU COVID Investigators
2. Additional Methods
3. Table 2 NAT score
4. Figure 1 Proportional distribution of cells in bronchoalveolar lavage fluid from patients with severe SARS-CoV-2 pneumonia.
5. Figure 2. Distribution of day of onset of ventilator-associated pneumonias
6. Figure 3. NAT score in response to bronchoalveolar lavage for all sampling times

Table 1. NU COVID Investigators

| Name | ORCID | email address | Affiliation 1 |
| --- | --- | --- | --- |
| A. Christine Argento | 0000-0002-3807-7690 | | Pulmonary and Critical Care Medicine |
| Ajay A. Wagh | 0000-0003-3402-0272 | | Pulmonary and Critical Care Medicine |
| Alexandra C. McQuattie-Pimental | 0000-0003-4028-9905 | | Pulmonary and Critical Care Medicine |
| Alexander V. Misharin | 0000-0003-2879-3789 | | Pulmonary and Critical Care Medicine |
| Alexis Rose Wolfe | 0000-0001-9321-892X | | Pulmonary and Critical Care Medicine |
| Alvaro Donayre | 0000-0003-1877-0655 | | Pulmonary and Critical Care Medicine |
| Ankit Bharat | 0000-0002-1248-0457 | | Thoracic Surgery |
| Anna E. Pawlowski | 0000-0002-9047-6437 | | Northwestern Medicine Enterprise Data Warehouse |
| Anne R. Levenson | 0000-0002-7587-1885 | | Pulmonary and Critical Care Medicine |
| Anthony M. Joudi | 0000-0003-1232-3256 | | Pulmonary and Critical Care Medicine |
| Benjamin D. Singer | 0000-0001-5775-8427 | | Pulmonary and Critical Care Medicine |
| Betty Tran | 0000-0001-6794-7088 | | Pulmonary and Critical Care Medicine |
| Catherine A. Gao | 0000-0001-5576-3943 | | Pulmonary and Critical Care Medicine |
| Chao Qi | 0000-0003-0741-3070 | | Department of Pathology |
| Chiagozie O. Pickens | 0000-0002-3400-9228 | | Pulmonary and Critical Care Medicine |
| Chitaru Kurihara | 0000-0003-3536-4675 | | Thoracic Surgery |
| Clara J Schroedl | 0000-0003-3280-4997 | | Pulmonary and Critical Care Medicine |
| Daniel Meza | 0000-0001-8154-1295 | | Pulmonary and Critical Care Medicine |
| Daniel Schneider | 0000-0002-3523-745X | | Northwestern Medicine Enterprise Data Warehouse |
| David A. Kidd | 0000-0003-0411-9511 | | Pulmonary and Critical Care Medicine |
| David D. Odell | 0000-0002-0121-4831 | | Thoracic Surgery |
| David W. Kamp | 0000-0001-5796-1206 | | Pulmonary and Critical Care Medicine |
| Elizabeth S. Malsin | 0000-0002-3294-6752 | | Pulmonary and Critical Care Medicine |
| Emily M. Leibenguth | 0000-0001-7809-7947 | | Pulmonary and Critical Care Medicine |
| Eric P. Cantey | 0000-0002-7065-7169 | | Cardiology |
| Gabrielle Y. Liu | 0000-0003-2049-423 | | Pulmonary and Critical Care Medicine |
| GR Scott Budinger | 0000-0002-3114-5208 | | Pulmonary and Critical Care Medicine |
| Helen K. Donnelly | 0000-0001-8827-8581 | | Pulmonary and Critical Care Medicine |
| Isaac A. Goldberg | 0000-0002-7178-9729 | | Pulmonary and Critical Care Medicine |
| Jacob I. Sznajder | 0000-0002-2089-4764 | | Pulmonary and Critical Care Medicine |
| Jacqueline M. Kruser | 0000-0003-3258-1869 | | Pulmonary and Critical Care Medicine |
| James M. Walter | 0000-0001-7428-3101 | | Pulmonary and Critical Care Medicine |
| Jane E. Dematte | 0000-0003-4907-9525 | | Pulmonary and Critical Care Medicine |
| Jason M. Arnold | 0000-0001-8687-0063 | | Department of Medicine |
| John Coleman | 0000-0003-1246-3814 | | Pulmonary and Critical Care Medicine |
| Joseph Isaac Bailey | 0000-0002-4225-3108 | | Pulmonary and Critical Care Medicine |
| Joseph S. Deters | 0000-0002-3522-3824 | | Pulmonary and Critical Care Medicine |
| Justin A. Fiala | 0000-0001-5196-7076 | | Pulmonary and Critical Care Medicine |
| Katharine Secunda | 0000-0003-0310-5783 | | Pulmonary and Critical Care Medicine |
| Kaitlyn Vitale | 0000-0002-8679-6200 | | Pulmonary and Critical Care Medicine |
| Khalilah L. Gates | 0000-0002-5965-8136 | | Pulmonary and Critical Care Medicine |
| Kristy Todd | 0000-0001-8210-8522 | | Pulmonary and Critical Care Medicine |
| Lindsey D. Gradone | 0000-0001-8145-2997 | | Pulmonary and Critical Care Medicine |
| Lindsey N. Textor | 0000-0001-7699-9429 | | Pulmonary and Critical Care Medicine |
| Lisa F. Wolfe | 0000-0001-9044-4147 | | Pulmonary and Critical Care Medicine |
| Lorenzo L. Pesce | 0000-0002-8015-4653 | | Thoracic Surgery |
| Luisa Morales-Nebreda* | 0000-0002-7126-0518 | | Pulmonary and Critical Care Medicine |
| Madeline L Rosenbaum | 0000-0001-9866-9850 | | Pulmonary and Critical Care Medicine |
| Manu Jain | 0000-0003-1534-5629 | | Pulmonary and Critical Care Medicine |
| Marc A. Sala | 0000-0002-2900-0595 | | Pulmonary and Critical Care Medicine |
| Mary Carns | 0000-0002-5063-156X | | Pulmonary and Critical Care Medicine |
| Marysa V. Leyla | [0000-0002-6583-5389](https://orcid.org/0000-0002-6583-5389) | | Cardiology |
| Mengjia Kang | 0000-0002-1679-9473 | | Pulmonary and Critical Care Medicine |
| Michael J. Alexander | 0000-0001-6271-3426 | | Pulmonary and Critical Care Medicine |
| Michael J. Cuttica | 0000-0002-0084-8760 | | Pulmonary and Critical Care Medicine |
| Michelle Hinsch Prickett | 0000-0002-1119-0079 | | Pulmonary and Critical Care Medicine |
| Natalie Jensma | 0000-0002-2046-5070 | | Hospital Medicine |
| Nicole Borkowski | 0000-0001-8650-6456 | | Pulmonary and Critical Care Medicine |
| Nikolay S. Markov* | 0000-0002-3659-4387 | | Pulmonary and Critical Care Medicine |
| Orlyn R. Rivas | 0000-0001-6938-4020 | | Department of Medicine |
| Paul A. Reyfman | 0000-0002-6435-6001 | | Pulmonary and Critical Care Medicine |
| Peter H. S. Sporn | 0000-0002-1006-9437 | | Pulmonary and Critical Care Medicine |
| Prasanth Nannapaneni | 0000-0001-8080-3214 | | Northwestern Medicine Enterprise Data Warehouse |
| Rachel B. Kadar | 0000-0002-2583-3637 | | Critical Care Medicine, Anesthesiology |
| Rachel M. Kaplan | 0000-0003-0038-9231 | | Cardiology |
| Rade Tomic | 0000-0003-2394-1287 | | Pulmonary and Critical Care Medicine |
| Radhika Patel | 0000-0002-1255-5818 | | Pulmonary and Critical Care Medicine |
| Rafael Garza-Castillon | 0000-0003-4970-6584 | | Thoracic Surgery |
| Ravi Kalhan | 0000-0003-2443-0876 | | Pulmonary and Critical Care Medicine |
| Richard G. Wunderink | 0000-0002-8527-4195 | | Pulmonary and Critical Care Medicine |
| Rogan A. Grant* | 0000-0003-0655-0882 | | Pulmonary and Critical Care Medicine |
| Romy Lawrence | 0000-0002-6578-8519 | | Pulmonary and Critical Care Medicine |
| Ruben J. Mylvaganam | 0000-0002-7203-7609 | | Pulmonary and Critical Care Medicine |
| Sean B. Smith | 0000-0001-8796-3966 | | Pulmonary and Critical Care Medicine |
| Samuel S. Kim | 0000-0001-8889-478X | | Thoracic Surgery |
| Sanket Thakkar | 0000-0002-0026-6533 | | Thoracic Surgery |
| SeungHye Han | 0000-0001-5625-6337 | | Pulmonary and Critical Care Medicine |
| Sharon R. Rosenberg | 0000-0002-0260-2545 | | Pulmonary and Critical Care Medicine |
| Susan R. Russell | 0000-0003-3006-1018 | | Pulmonary and Critical Care Medicine |
| Sydney M. Hyder | 0000-0002-6395-9062 | | Pulmonary and Critical Care Medicine |
| Taylor A. Poor | 0000-0002-2191-5037 | | Pulmonary and Critical Care Medicine |
| Theresa A. Lombardo | 0000-0002-7468-7541 | | Pulmonary and Critical Care Medicine |
| Zasu M. Klug | 0000-0003-3483-396X | | Pulmonary and Critical Care Medicine |

**Additional Methods**

**Patient Cohort**

The first SARS-CoV-2 positive patients were admitted to the medical ICU in early March 2020. Up to 67 ventilated patients per day were cared for in four ICUs repurposed to treat patients with COVID-19-induced critical illness as part of the initial pandemic surge management. All patients were cared for in closed rooms that met engineering standards for negative pressure.

We accessed a COVID-19 datamart created within the Northwestern Medicine Enterprise Data Warehouse (NMEDW) for clinical data collection. Patients were included in the cohort if they had a positive COVID-19 electronic flag and required mechanical ventilation after March 1, 2020. The electronic flag indicated patients who had a positive polymerase chain reaction (PCR) assay for SARS-CoV-2 from a nasopharyngeal swab or BAL fluid during the index hospitalization or patients who tested positive at another facility or as an outpatient before the index hospitalization.

Clinical management, including biomarker testing, intubation thresholds, ventilator care, airway therapy, sedation and thromboprophylaxis, was guided by standardized protocols developed by a multidisciplinary panel led by critical care physicians that included experts from infectious disease, interventional pulmonary, hospital medicine, anesthesia, neurology critical care and ethics. These protocols were updated continuously over the course of the pandemic and were available electronically to all providers via intranet.

Demographics, clinical laboratory values, and medications were extracted from the NMEDW. The electronic medical record was selectively reviewed for data that could not be electronically extracted, particularly for patients transferred from other institutions. Enrollment in placebo-controlled trials of remdesivir or sarilumab was extracted from medication records by physician chart review. The Northwestern University Institutional Review Board (STU00212283) approved this study.

**Bronchoscopic sampling**

A BAL early after endotracheal intubation is our center’s standard of clinical care for patients with suspected pneumonia. We have reported that sampling of the alveolar space by BAL in patients with pneumonia is associated with low rates of complications.^1^ While BAL is typically obtained in our center using non-bronchoscopic sampling by respiratory therapists, this procedure was considered a high-risk aerosol-generating procedure in patients with COVID-19. We therefore modified our standard bronchoscopic BAL technique for ventilated patients in order to minimize aerosol generation. ^2^ Nurses and respiratory therapists were encouraged to leave the room during the bronchoscopy.

BAL was obtained at the discretion of the clinical team and results were used to target or discontinue empirical antibiotic therapy. All bronchoscopies were performed by pulmonary and critical care physicians using a disposable bronchoscope (aScope™ 4 Broncho endoscope, Ambu A/S, Denmark). The recommended installation volume was 120 ml and return from the initial 30-ml aliquot was discarded. The majority of BALs were performed for suspicion of bacterial superinfection; a few were performed therapeutically to remove secretions with fluid obtained for diagnostic studies.^3^ No bronchoscopist developed COVID-19 over the course of our study.^2^

**Analysis of BAL fluid**

BAL fluid cell counts were obtained via a hemocytometer and differential cell counts were performed on a cytocentrifuged slide with staining (Wright-Giemsa or May-Grunwald-Giemsa) with enumeration of at least 100 cells. At the onset of the pandemic, BAL fluid cell counts were temporarily unavailable until the clinical laboratory received appropriate safety certification. Before the pandemic, we had internally validated the BioFire FilmArray RP2 Pneumonia Panel,^4^ a rapid multiplex PCR assay that identifies 33 viral and bacterial pathogens and resistance genes (See supplementary materials). This panel received institutional approval for routine testing just before admission of the first ICU patient with severe SARS-CoV-2 pneumonia. This assay was used for most BALs but limited supplies during the pandemic prevented its use for all samples. When the BioFire Pneumonia Panel was unavailable, BAL samples were analyzed for respiratory viruses using a multiplex PCR array (BioFire Respiratory Panel) and a locally validated methicillin-resistant *S. aureus* (MRSA) PCR (Cepheid SSTI).^5^ All samples underwent quantitative bacterial cultures using a calibrated loop technique and fungal cultures. The lower limit of detection for quantitative cultures was 10 colony-forming units (cfus)/ml. A BAL amylase assay was used to assess for the degree of aspiration.^6^ We recorded the proportion of neutrophils in the fluid samples as the proportion of neutrophils in BAL fluid <50% has a high negative predictive value for presence of bacterial pneumonia.^1,7^ We reviewed patients in our database of patients with non-COVID-19 pneumonia to define the upper 95% confidence interval for the percentage lymphocytes in BAL fluid as 10%.^8^

**Definitions**

We defined early bacterial superinfection as detection by culture or multiplex PCR of a respiratory pathogen known to cause pneumonia in addition to SARS-CoV-2 on a BAL specimen collected within 48 hours after intubation for COVID-19 pneumonia. We defined VAP as detection of a new respiratory pathogen in BAL fluid obtained more than 48 hours after intubation. Recurrent VAP was defined as culture/PCR positivity for the same pathogen with an interval negative BAL result. Persistence of VAP was defined as the same pathogen in BAL culture or PCR. Persistent VAP was not considered a new episode unless an interval BAL was negative and a treatment course had been concluded. For both initial superinfection and VAP, pneumonia was defined as detection of a respiratory pathogen known to cause pneumonia by PCR, quantitative bacterial culture or fungal culture, or the detection of antigens for *Legionella pneumophila* serogroup 1 or *Streptococcus pneumoniae* antigen in a urine specimen. To avoid excluding potential causes of pneumonia, we considered a quantitative culture positive if organisms were detected at 10 cfu/ml or higher.

Beta-lactam antibiotics are considered the backbone of pneumonia treatment.^9^ We therefore categorized the antibiotic resistance of bacterial pathogens according to the recently described category of Difficult-to-Treat Resistance, which is based on resistance to beta-lactam antibiotics.^10^

**Narrow Antibiotic Treatment Score**

We developed a scoring system, Narrow-spectrum Antibiotic Treatment (NAT) score, that provides a relative daily measure of the spectrum of antimicrobial coverage provided by the antibiotic or antibiotic combinations administered on each hospital day (Supplemental Table 2). The NAT score was developed in anticipation of a randomized controlled trial on the benefit of a rapid diagnostic test for severe community-acquired pneumonia.^11^ The baseline was therefore set as combination therapy with ceftriaxone and either azithromycin or a respiratory fluoroquinolone, according to the ATS/IDSA guidelines.^12^ This regimen was scored as 0 in order to be able to calculate either narrower or broader spectrum treatment. Use of monotherapy with these agents would therefore result in a -1 score for that day whereas broader-spectrum antibiotics would give a positive score.

Every antibiotic given during the calendar day was totaled for the day. **No antibiotic therapy would result in a -2 score for the day**.

Supplemental Table 2. Daily Narrow-spectrum Antibiotic Treatment (NAT) Score

| **Regimen** | **Score** |  |
| --- | --- | --- |
|  | Combination | Monotherapy |
| **Ceftriaxone** | 0 | -1 |
| **Azithromycin** | 0 | -1 |
| **Respiratory fluoroquinolone**  **(levofloxacin, moxifloxacin, etc)** | 0* | -1 |
| **Cefazolin** | 0 | -1 |
| **Piperacillin/tazobactam** | +1 | +1 |
| **Vancomycin** | +1 | +1 |
| **Linezolid** | +1 | -1 |
| **Clindamycin** | +1 | -1 |
| **Cefipime** | +1 | +1 |
| **Cefpodoxime** | +1 | -1 |
| **Ampicillin/sulbactam** | +1 | -1 |
| **Amoxicillin/clavulanic acid** | +1 | -1 |
| **Meropenem** | +2 | +2 |
| **Ceftolozane/tazobactam** | +3 | +3 |
| **Ceftazidime/avibactam** | +3 | +3 |
| **Aminoglycoside** | +1 | N/A |
| **Trimethoprim/sulfamethoxazole** | +1 | -1 |
| **No antibiotic** | N/A | -2 |

Supplemental Figure Legends

Supplemental Figure 1. Proportional distribution of cells in bronchoalveolar lavage fluid from patients with severe SARS-CoV-2 pneumonia.

Supplemental Figure 2. Distribution of day of onset of ventilator-associated pneumonia

Supplemental Figure 3. NAT score in response to bronchoalveolar lavage for all sampling times
