## Supplementary figures and images for "Bacterial superinfection pneumonia in SARS-CoV-2 respiratory failure"

### Supplementary Figure 1

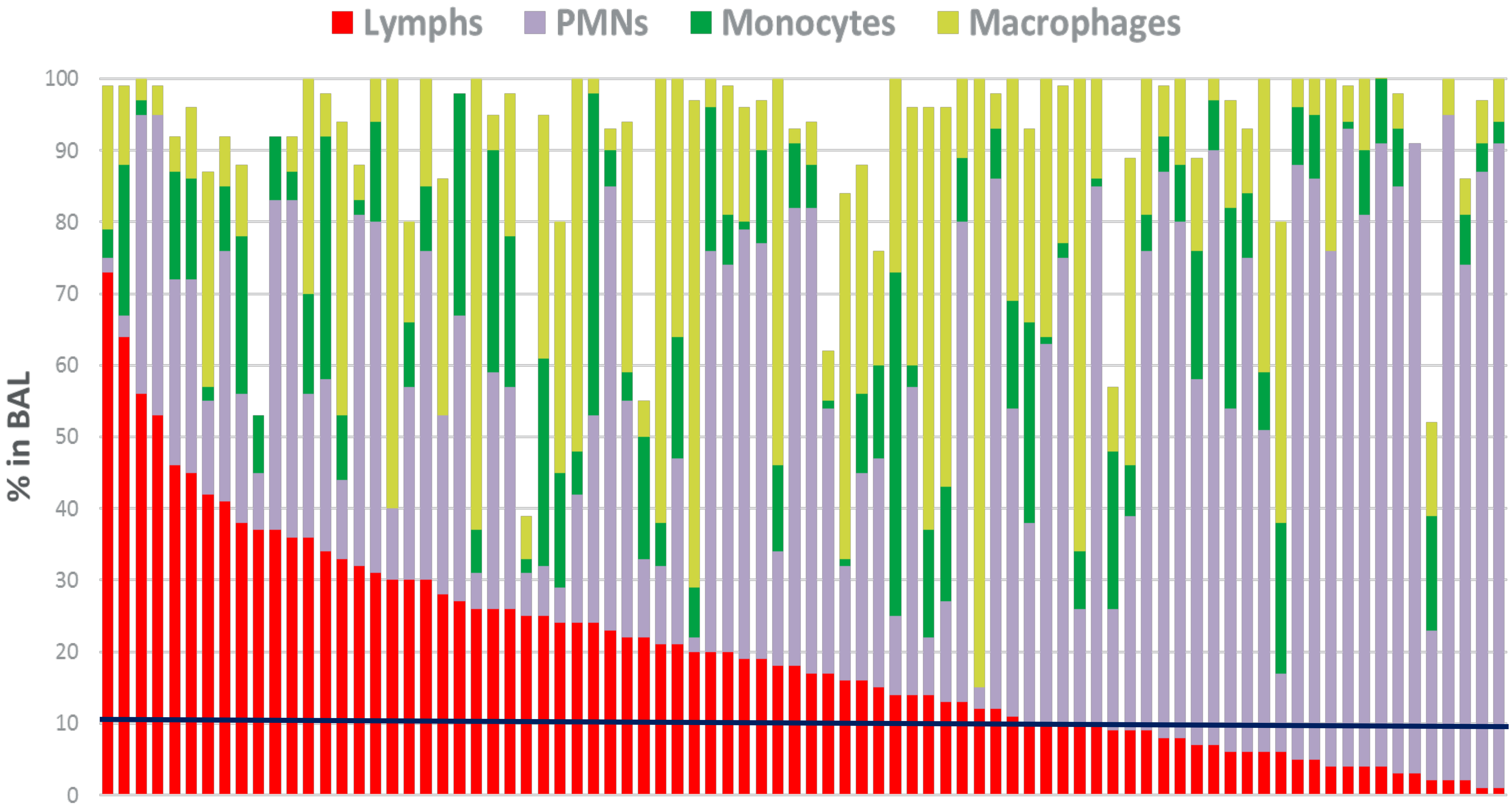

### Supplementary Figure 2

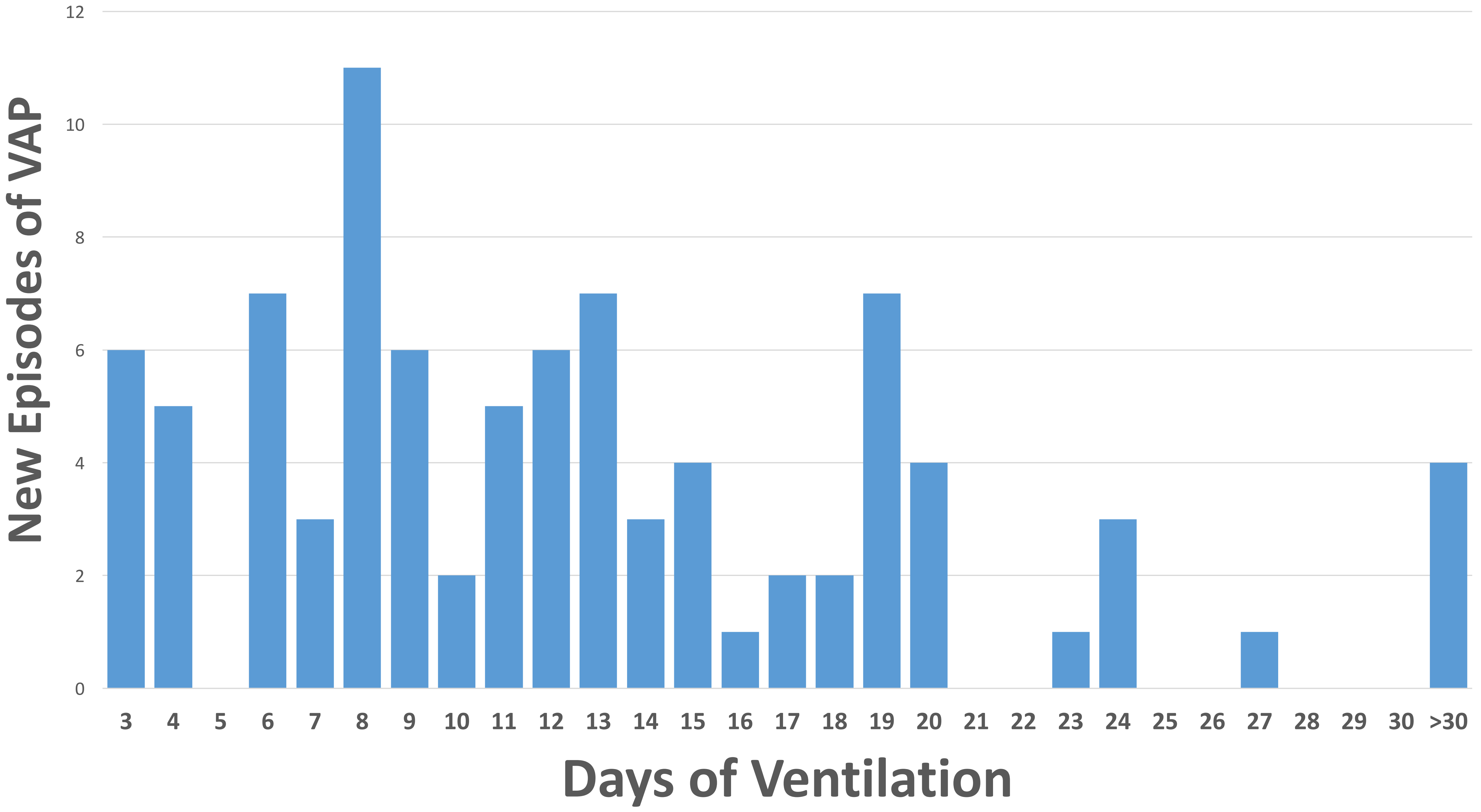

### Supplementary Figure 3

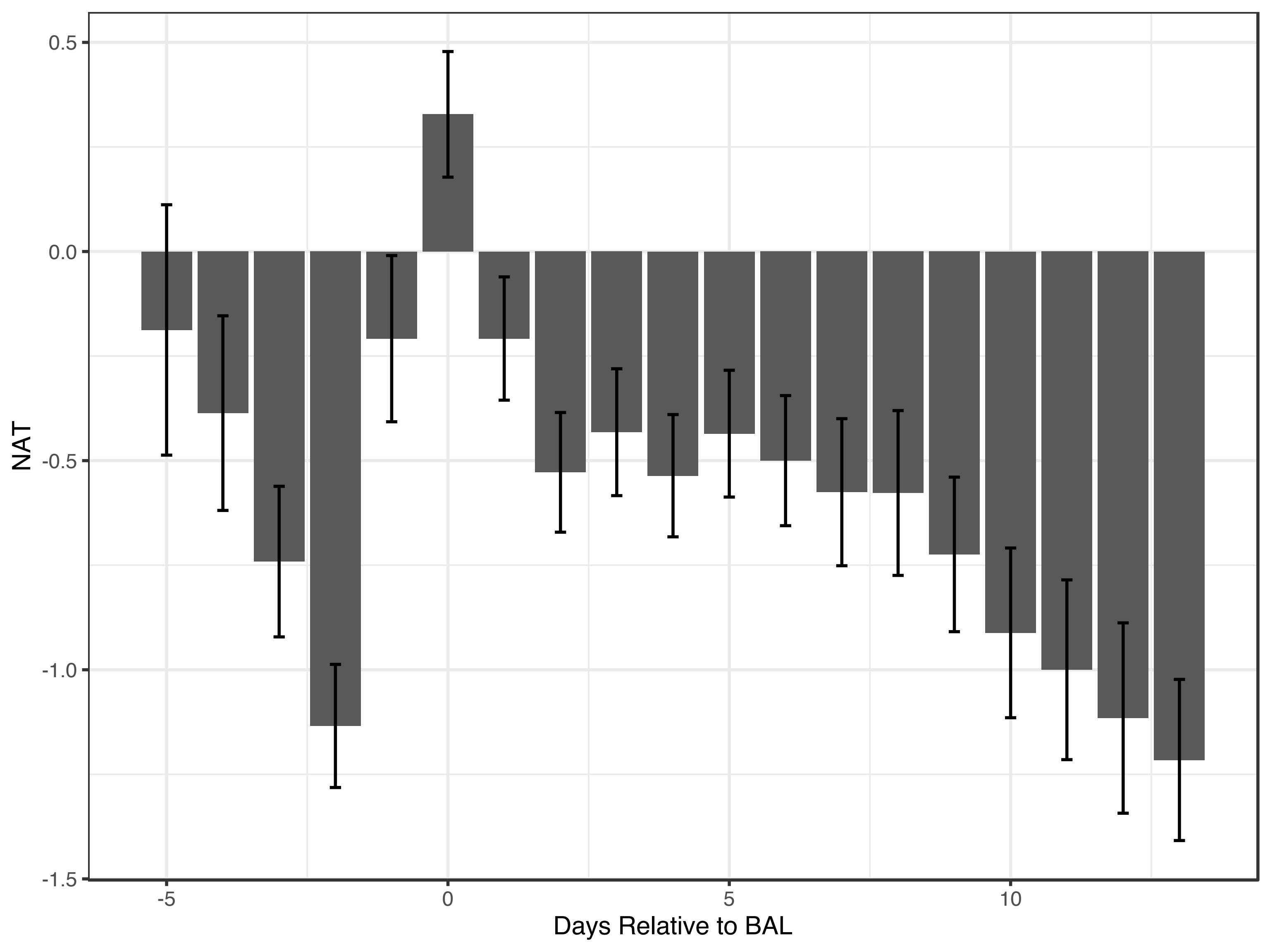
